# Home-based binocular serious games in virtual reality to treat visual acuity and stereovision in residual amblyopia: AMBER study

**DOI:** 10.64898/2026.06.12.26355255

**Authors:** Aurelia Aurilia, Nour-Louise Martin, Cristina Simon-Martinez, Maria-Paraskevi Antoniou, Walid Bouthour, Daphne Bavelier, Benjamin T Backus, Brian Dornbos, James J Blaha, Martina Kropp, Henning Müller, Micah M. Murray, Gabriele Thumann, Heimo Steffen, Pawel J Matusz

## Abstract

**Objectives:** Amblyopia is a pediatric visual disorder traditionally treated by patching the fellow eye, though many patients retain residual amblyopia post-treatment. Increasing evidence suggests that visual plasticity allows treatment beyond the classical therapeutic window. AMBER evaluated the efficacy of binocular serious games in virtual reality (VR) in residual amblyopia.

**Methods and Analysis:** The monocentric, prospective, randomized, crossover trial (reported as case series) included 14 anisometropic, strabismic, or mixed residual amblyopia patients (6-35 years; 5 children, 9 adults). Participants underwent two 2-month intervention phases: optical correction (standard care) and standard care plus VR games (2.5 h/week), each with a 2-month follow-up. Best-corrected visual acuity (BCVA), stereoacuity, and reading speed were assessed (5 timepoints) using the Sloan and Landolt charts, the Titmus, TNO, Lang II, Asteroid, and Mnread tests. Compliance and adverse events (AE) were recorded.

**Results:** VR training improved BCVA in 10 amblyopic eyes (Landolt and Sloan), with more pronounced effects in anisometropic patients. Six patients showed improved stereoacuity (Titmus; 4x mixed, 1x anisometropic, 1x strabismic amblyopia), persistent only in children (1x strabismic, 1x mixed amblyopia). Four improvements were observed with TNO (1x), Lang II (1x), Asteroid (0x), and MNread (1x). Despite positive trends, when comparing results of individual patients, between both eyes, and with standard treatment, consistency of improvements cannot be conclusively demonstrated. One non-severe AE (dizziness) was reported.

**Conclusions:** Following individual cases, VR training improved BCVA and stereoacuity, particularly in children and patients with high compliance. However, considering the cohort as a whole, consistency of effects has to be confirmed in larger groups. Thus, the methodologically sophisticated AMBER study revealed differences in VR treatment efficacy between amblyopia types, children/adults, endpoints and tests, offering precious data for the design of meaningful future studies. It shows that neurovisual plasticity gauged by VR-games offers safe, engaging treatment options for residual amblyopia.

**What is already known on this topic:**

- Binocular treatment approaches have shown promise in efficiently treating amblyopia for which the standard treatment, patching, is effective but is limited by factors such as poor compliance and a narrow treatment window.

**What this study adds:**

- The study investigated the benefits of binocular serious games in virtual reality for both children and adults with residual amblyopia and compared various tests to assess outcomes. This comprehensive approach allowed for a comparison of treatment efficacy across different age groups and types of amblyopia, as well as an evaluation of the suitability of the various tests for determining treatment outcomes.

**How this study might affect research, practice or policy:**

- The findings support the hypothesis that binocular training can effectively treat amblyopia demonstrating highest safety and benefits even beyond the classical treatment window, and in residual amblyopia, which could affect clinical practice of ophthalmologists who offer such therapies, at least as a supplement to standard treatment.
- Assessment of various tests will help uniformize future studies, thus, increase studies’ comparability and, finally, allow conclusive analyses and formulation of treatment recommendations.

## 1. Introduction

The developmental visual disorder amblyopia is characterized by reduced visual acuity (VA) of one eye that cannot be improved by optical correction^1^. It is mainly caused by anisometropia or strabismus affecting about 3.4% of the population^1^. Considered a monocular disorder, standard treatment relies on occlusion of the fellow eye with proven efficacy in children from 3-6 years of age^2^. However, patching is limited by poor compliance and does not address binocular deficits such as impaired stereovision, reading speed, attention and fine motor skills^3^, and many patients are left with residual amblyopia^4^.

Recent studies demonstrated that vision can improve beyond the age of 6 years through perceptual learning and monocular video game-based interventions^5–7^. In parallel, amblyopia has increasingly been defined as a binocular disorder characterized by interocular suppression and impaired binocular fusion, leading to the development of dichoptic approaches, in which each eye is presented with complementary stimuli to promote binocular cooperation^8^. Vedamurthy *et al*. demonstrated reduced suppression, VA and stereovision improvements in adults using a dichoptic virtual reality (VR) game; though, the mechanisms remained unclear due to combined attentional and dichoptic effects^9^. Later, they evaluated binocular 3D interventions in amblyopic stereoblind adults and reported stereopsis improvements using a bug squashing game^10^. Gambacorta *et al*. evaluated a binocular video game in children with anisometropic and strabismic amblyopia (7-17 years), reporting greater VA and stereopsis improvements compared with patching, although efficacy remained limited, likely due to high attentional demands^11^. Molina-Martín *et al*. reported contrast sensitivity improvements with an immersive dichoptic VR program in anisometropic children (8-14 years)^12^. While promising, small sample size and large differences in patients’ cohorts (age, amblyopia type), VR interventions, and treatment duration limit comparability and reproducibility of these results. Some interventions combined multiple methods, thus leaving therapeutic mechanisms unclear. These aspects made it difficult to assess clinical relevance of binocular VR approaches in amblyopia. The findings call for rigorous approaches, comparing different phenotypes, games, ages, treatment duration, and designs adapted to children’s capacities.

Differences in measurement methods of best corrected VA (BCVA), may also influence outcomes. Previous studies have used either Sloan or Landolt charts, but rarely both. Combining both measures in the present study provided a more reliable evaluation of visual improvement. Similarly, stereoacuity has typically been assessed using one test (mostly Randot or TNO), which may not capture the full range of binocular acuity, potentially increasing the risk of false positives results.

Present prospective, single-center, randomized crossover clinical trial (RCT) evaluated the effectiveness of binocular stimulation using eight complementary serious VR-based games as an adjunct to optical correction (OA) in patients with residual amblyopia of age beyond the therapeutic window (protocol paper published^13^). The comparison of multiple BCVA and stereopsis tests aimed at identifying differences and finding the most appropriate (combination of) methods to be used. We hypothesized that VR-based games would improve BCVA, stereovision, reading, attentional and motor skills in patients with residual amblyopia, regardless of age. The RCT demonstrated benefits in three outcomes tested, in some patients. The efficacy of VR in amblyopia varied depending on age, amblyopia type, compliance, and the test measuring the outcome. The consistency of the effects has to be confirmed in larger groups. Although the study was designed as an RCT, the limited sample size and heterogeneity of the study population led us to interpret the findings as a case series.

## 2. Materials and Methods

### 2.1. Study design and ethics statement

AMBER was a prospective, blinded, monocentric, cross-over RCT, conducted in accordance with Declaration of Helsinki & approved by the Ethical Cantonal commission of Geneva (N° 2021-D0090, approved 8.11.22); the limited number and heterogeneity of enrolled patients led us report the results as case series. The protocol followed the SPIRIT (Standard Protocol Elements: Recommendations for Interventional Trials) Statement and examinations were performed at the Department of Ophthalmology, University Hospitals of Geneva (HUG Ophthalmology).

Patients (nonblinded) completed two 2-month-long interventions in a randomized order: OA (optical correction alone) (patients wore glasses with prescribed correction), and VR (patients played binocular serious video games in addition to OA). Randomization was stratified according to amblyopia type (anisometropic, strabismic or mixed), using REDCap. Each phase was followed by a 2-month follow-up, resulting in a study duration of 8 months/participant. Two certified, orthoptists performed ocular examinations during 5 visits scheduled per patient; another team member assessed non-ocular outcomes. All (and other two members, analyzing the data) were blinded. Visits took place at baseline (T1, screening), after the first intervention (T2, short-term [ST]), 2 months after the end of the first intervention (T3, follow-up 1, long-term [LT] and screening for the second intervention), after the second intervention (T4, [ST]), and after the second follow-up (T5, [LT]) (Fig 1).

**Figure 1.**
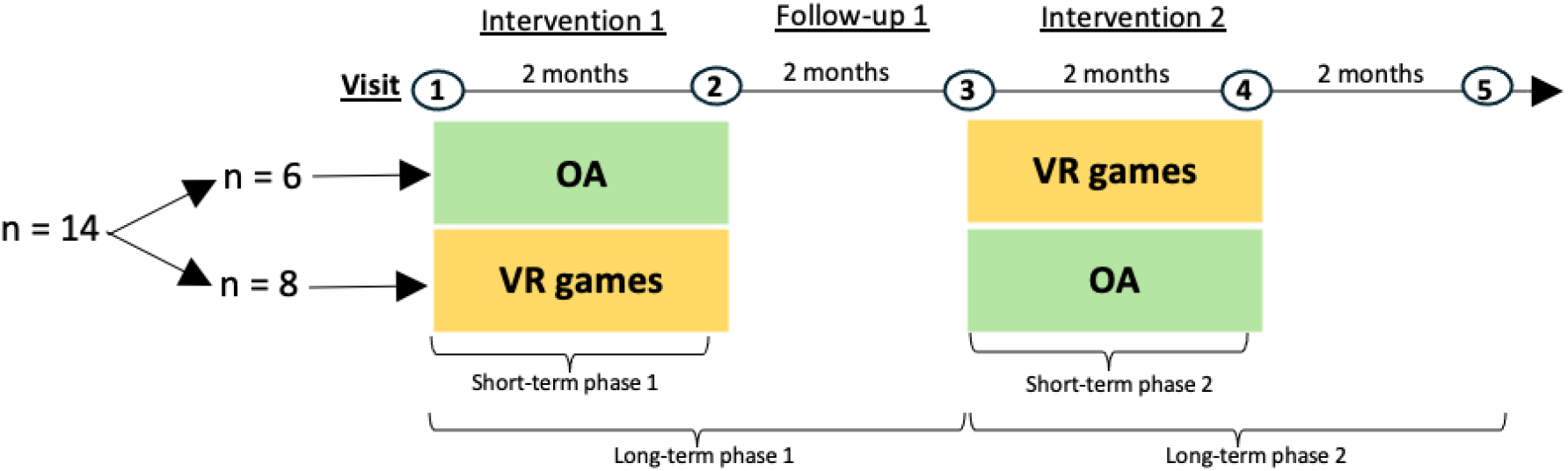
The AMBER cross-over design.

### 2.2. Participants

Eligible participants were aged 6-35 years with residual amblyopia (anisometropic, strabismic or mixed), defined as BCVA < 20/20 in the amblyopic eye (AmE) and an interocular difference of ≥2 lines with refractive correction. BCVA had to be stable across at least 2 consecutive assessments obtained ≥6 months after the last treatment. Exclusion criteria included: untreated or recently diagnosed amblyopia, atropine use within 3 months prior to study enrollment, prior eye surgery (except strabismus surgery), strabismus exceeding 20D or with large eccentric fixation, any additional uncorrected visual, ocular, neurological, developmental, psychological sensorimotor or auditory disorder.

Nineteen patients were recruited between 05/2022 and 12/2024 at HUG Opthalmology. Two patients did not meet inclusion criteria, three dropped out after T1. Ultimately, 14 patients were evaluated (data of all are reported here), with 13 completing both interventions. Six participants were allocated to the VR→OA sequence and eight to the OA→VR sequence. Demographic and clinical characteristics (gender, age, handedness, dominant eye, previous eye surgery, amblyopia type) were collected.

### 2.3. Virtual reality games arm

The VR arm involved eight binocular serious games embedded in a VR headset, allowing home-based training after an introduction at the clinic (Vivid Vision, San Francisco, USA). Games were played, while wearing prescribed glasses, for 30 min/day limited to this maximum duration by automatic locking, 5 days/week, over 2 months. The software adjusted interocular contrast based on performance, gradually reducing the difference between the AmE and the fellow eye (FE) to increase task difficulty while maintaining feasibility, to equalize binocular input, and to improve depth perception. Games were organized following the hierarchy of binocular vision, from simultaneous perception and anti-suppression to fusion and vergence, and finally stereopsis. Each session included anti-suppression followed by stereopsis tasks, while vergence training was introduced once a minimum stereoacuity threshold was reached (assessed every five sessions). Games targeting anti-suppression followed, when using dichoptic stimuli:

- *Breaker*: In this first game of each session a paddle (seen by FE) is used to hit a ball (seen by AmE) to destroy a wall of bricks (seen by both eyes);
- *Ring Runner:* a spaceship (seen by FE) needs to fly through rings (seen by AmE);
- *Hoopie:* a basketball (both eyes), with X or O markers (seen by AmE) needs to be caught by moving the hoop;
- *Pepper Picker:* peppers (seen by AmE) need to be picked up, appearing at different depths and heights (seen by FE).

Stereopsis training included:

- *Bubbles*: floating balls at different depths are popped. A “scaffolding” feature allows motion parallax cues that is gradually reduced to zero as the score improves. The game is included in every session and if minimum performance is not met, no other games are played afterwards;
- *Bullseye*: targets positioned at various planes and depths have to be shot.

Games targeting vergence:

- *Barnyard Bounce*: an animal has to jump to higher platforms.

### 2.4. Primary outcome

Primary outcome was BCVA measured by the Sloan and the Landolt charts in both eyes at 5 m. Values were documented in decimal form before being converted to logMAR. Changes in BCVA were assessed in ST and LT as the difference in logMAR values between timepoints. ST change was defined as T2-T1 (1^st^ arm) or T4-T3 (2^nd^ arm), while LT change was defined as T3-T1 (1^st^ arm) or T5-T1 (2^nd^ arm). Percentage changes were calculated relative to baseline (T1) using the formula (Tx-T1)/T1.

### 2.5. Secondary outcomes

Stereovision, reading speed, and compliance were considered secondary outcomes.

#### 2.5.1 Stereoacuity

Stereoacuity was measured using the Titmus, TNO and Lang II tests and a tablet-based random-dot stereotest^14^. Variations over time were evaluated in arcsec and expressed as percentage from baseline (for ST and LT).

##### Titmus

Includes three tests - the Fly, Circles, Animals^15^. Only if the patient has stereovision, the fly’s wings appear in three-dimensional (3D) form. Patients unable to perceive the wings were assigned a value of 3,600 arcsec and classified as stereoblind. The latter quantify stereoacuity as Circles: 800-40 arcsec, Animals: 400-100 arcsec. Those who perceived the fly but could not identify any circle or animal were assigned a value of 3,552 arcsec.

##### TNO

Consists of six plates. Plates I-III screen for stereovision (threshold 1,980 arcsec). Plate IV detects interocular suppression, plates V and VI quantify stereoacuity from 480-60 arcsec.

##### Lang II

Includes three images – elephant, truck, moon – corresponding to 600, 400 and 200 arcsec, respectively, and one monocularly visible star. If only the star was seen, 1,800 arcsec were assigned.

##### Asteroid

Patients identify among four dynamic stereograms the one containing depth^14^. The difficulty level, i.e., the detectable depth, changes on a trial-by-trial basis according to the patient’s performance. Asteroid measures stereopsis along a continuous range. Every possible level within the measurable range (13-1,200 arcsec) is assessed. Asteroid does not contain monocular cues, so stereoblind patients (attributed 1,200 arcsec) cannot identify depth-containing stereograms.

#### 2.5.2. Reading speed

The MNRead test, requiring fluent reading, was performed with patients of ≥8 years of age and conducted on an iPad, separately for each eye, using the 2016 French electronic version. MNRead evaluates reading skill level in individuals with low vision, using short, standardized sentences of child-appropriate language. The text becomes increasing smaller across trials, while the complexity remains constant, ensuring that errors and speed are influenced solely by visual abilities. Recorded metrics were the smallest print size read without errors, the smallest print size read at maximum speed, the maximum reading speed measured in words per minute (wpm).

#### 2.5.3. Adverse events (AEs) and compliance

AEs were recorded by asking participants about disabling headaches, nausea, dizziness, or other. Patients or children’s parents documented daily AEs and the amount of time played in a phone-based app diary implemented on REDCap. The research team was notified immediately about any AE or if a patient did not play for >2 days.

VR compliance was defined as the percentage of total gameplay time out of the total 20 h. It was considered low with <33% (<7 h), medium 34-66% (7-13 h) or high between 67-100% (>13 h) of playing.

### 2.6. Statistical analysis

Target sample of 30 participants, powered for primary outcome^13^, was not reached due recruitment around the COVID19 pandemic. The small, heterogeneous sample led us analyze data descriptively (min, max, SD, SEM, range), reporting individual patient outcomes as detailed case studies. If not otherwise mentioned, data are reported as mean±SD, separately for adults and children, and 3 amblyopia subtypes.

## 3. Results

### 3.1. Sample characteristics

All 14 patients attended T1 and T2. At T3, 1 patient dropped out (n = 13), another before T4 (n = 12), and a third one by T5 (n = 11). Eight Patients were male (57%), 6 were female (43%), the age ranged 9-35 y (21.7±9.8 y); 5 were children aged between 8 and 12 (10.4±0.9 y), and 9 adults ranged from 21-35 (28.2±5.3 y). Three patients had diagnosis of anisometropic amblyopia, 5 strabismic (3 that had undergone strabismus correction surgery) and 6 mixed amblyopia. Twelve patients were right-handed and in 9 patients the right eye was the AmE. All BCVA and stereoacuity measures had 4 and 2 missing data, for VR and OA, respectively (12 of these ST, 24 LT).

### 3.2. Visual acuity (AmE)

#### 3.2.1 Landolt Chart

In the VR arm, 3 children (A1, S1, S3) improved at ST, with 1 maintaining gains at LT; 1 (M1) remained stable at ST and LT; and 1 (S2) deteriorated. Among adults, 5 (A2, A3, S5, M2, M5) improved at ST, with 2 maintaining gains at LT; 1 (M4) remained stable at ST but deteriorated at LT; and 2 (S4, M3) deteriorated at ST and further declined at LT (Table 2 & Fig 2).

**Figure 2.**
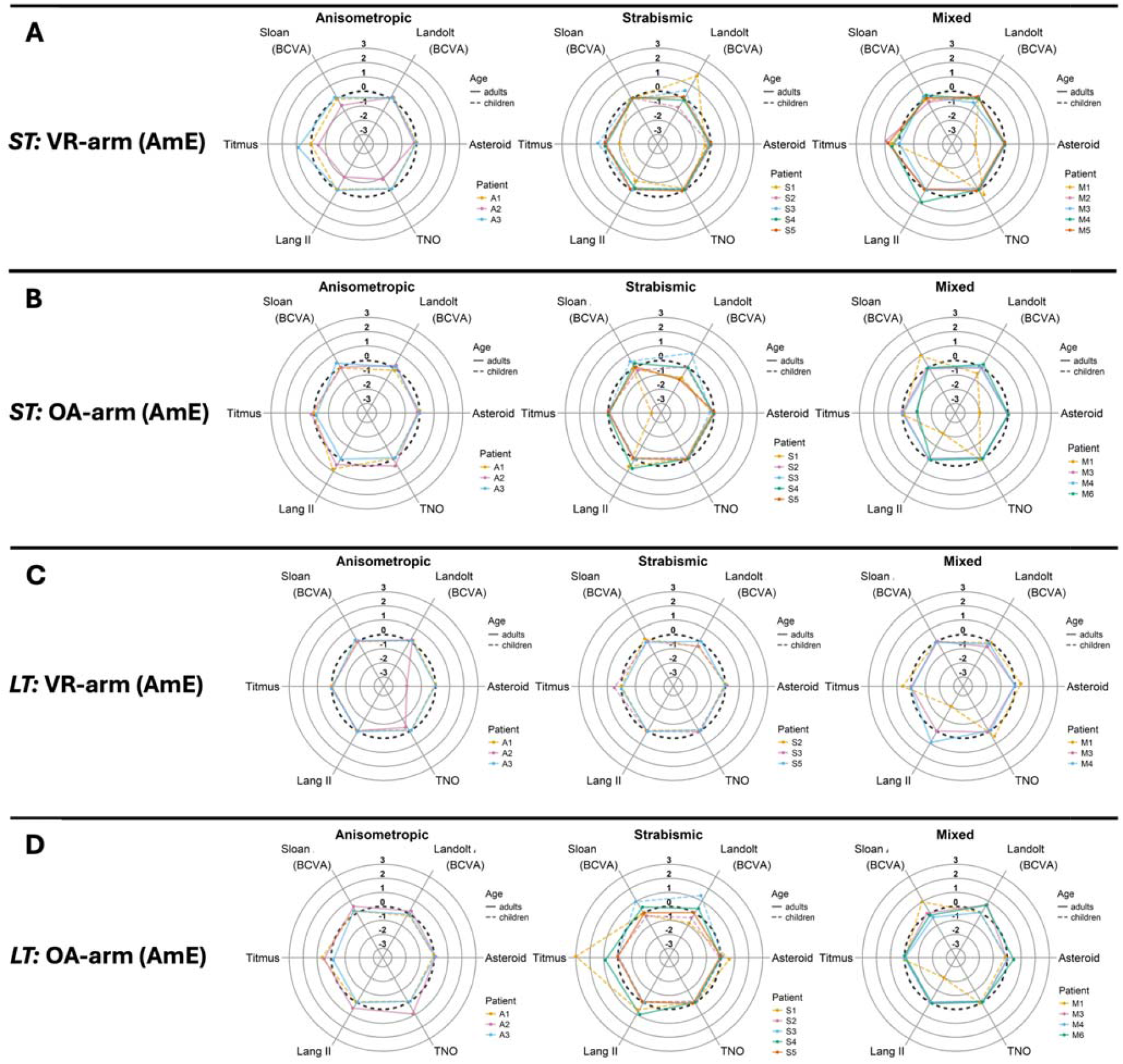
Radar plots showing the effects in BCVA and stereoacuity in the AmE at ST and LT, across VR and OA. Dotted reference line, indicates zero change from baseline; continous line, adults; dashed line, children.

In the OA arm, 1 child (S3) improved at ST and LT, 1 (S2) remained stable at ST, and 3 (A1, S1, M1) deteriorated at ST, with 2 maintaining this at LT. Among adults, 3 (A2, A3, M6) improved at ST, with 2 maintaining gains at LT; 4 (S4, M3, M4, M5) remained stable; and 1 (S5) deteriorated at ST but returned to baseline at LT (Table 2 & Fig 2).

#### 3.1.2. Sloan Chart

In the VR arm, all children remained stable at ST and LT except S2, who improved at LT. Among adults, 3 (A3, M3, M4) improved at ST, with 1 maintaining gains at LT; 3 (S4, S5, M5) remained stable, and 2 (A2, M2) deteriorated (Table 2 & Fig 2).

In the OA arm, 2 children (S3, M1) improved at ST maintaining gains at LT; 2 (A1, S1) remained stable; and 1 (S2) deteriorated at ST and LT. Among adults, 2 (A3, S4) improved at ST and LT, while 6 (A2, S5, M3, M4, M5, M6) remained stable (Table 2 & Fig 2).

### 3.2. Stereoacuity

#### 3.2.1 Titmus test

Ten patients showed measurable stereoacuity at baseline.

In the VR arm, 2 children (S3, M1) improved at ST and LT, 2 (A1, S2) remained stable, and 1 (S1) worsened. Among adults, 4 (A3, M2, M4, M5) improved at ST, but not LT; 3 (S4, S5, M3) remained stable and 1 (A2) declined.

**Table 1.** Patients’ demographics and compliance. ID, patients’ code; amblyopia types: A, anisometropia; S, strabismus; M, mixed; Age, years. The last two columns present compliance with the VR intervention as time elapsed (hours:minutes) and percentage of the fulfilled prescribed playing time (20 hours). *Note*: Patient age is expressed in ≥5-year age ranges, to avoid patient-identifying information.

| ID | Age<br>rang<br>e | Surgery | Type of strabismus | VR time (h:m) | Compliance |
| --- | --- | --- | --- | --- | --- |
| A1 | 8-12 | No | - | 3 : 33 | 18% |
| A2 | 21-25 | No | - | 15 : 33 | 78% |
| A3 | 31-35 | No | - | 17 : 48 | 89% |
| S1 | 8-12 | Strabismus correction | esotropia | 14 : 40 | 73% |
| S2 | 8-12 | No | exotropia | 6 : 52 | 34% |
| S3 | 8-12 | Strabismus correction | esotropia, hypotropia | 10 : 41 | 53% |
| S4 | 26-30 | No | esotropia | 19 : 58 | 100% |
| S5 | 31-35 | No | esotropia, hypotropia | 9 : 20 | 47% |
| M1 | 8-12 | No | exophoria (close & far) | 12 : 45 | 64% |
| M2 | 21-25 | No | exotropia | 29 : 01 | 100% |
| M3 | 21-25 | No | exotropia, hypotropia | 13 : 57 | 70% |
| M4 | 26-30 | No | esotropia | 8 : 02 | 40% |
| M5 | 31-35 | Strabismus correction | hypotropia | 19 : 29 | 97% |
| M6 | 31-35 | No | esotropia | 2 : 00 | 10% |

**Table 2.**
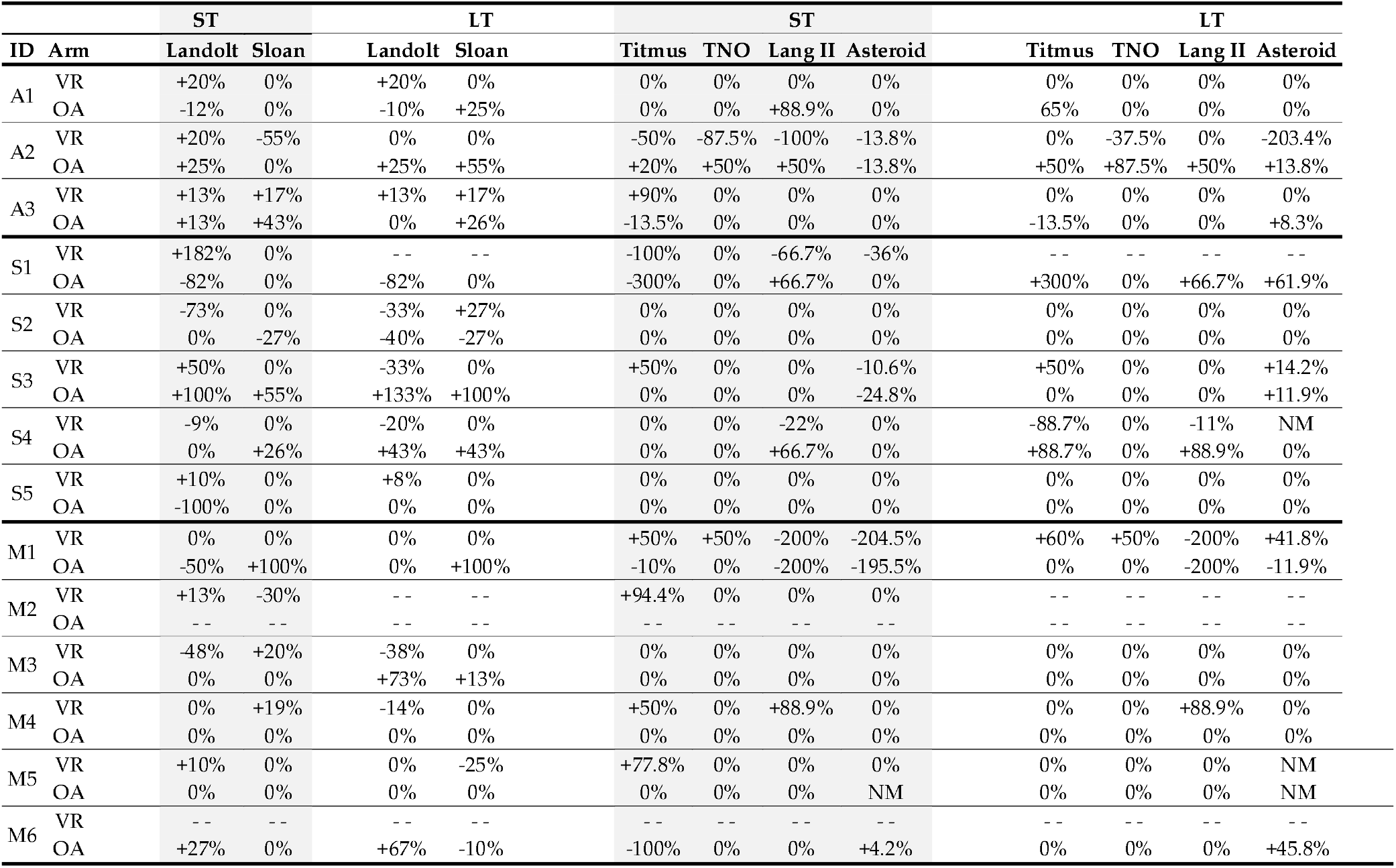
BCVA changes measured using Landolt and Sloan charts & stereoacuity measured using the Titmus, TNO, Lang II and Asteroid tests. Improvements are marked with a “+”, deteriorations with a “-” and stability with “0%”. The symbol “-- “ indicates that the patient dropped out. NM indicates that the measure was not taken.

|  |  | ST |  | LT |  | ST |  |  |  | LT |  |  |  |
| --- | --- | --- | --- | --- | --- | --- | --- | --- | --- | --- | --- | --- | --- |
| ID | Arm | Landolt | Sloan | Landolt | Sloan | Titmus | TNO | Lang II | Asteroid | Titmus | TNO | Lang II | Asteroid |
| A1 | VR | +20% | 0% | +20% | 0% | 0% | 0% | 0% | 0% | 0% | 0% | 0% | 0% |
|  | OA | -12% | 0% | -10% | +25% | 0% | 0% | +88.9% | 0% | 65% | 0% | 0% | 0% |
| A2 | VR | +20% | -55% | 0% | 0% | -50% | -87.5% | -100% | -13.8% | 0% | -37.5% | 0% | -203.4% |
|  | OA | +25% | 0% | +25% | +55% | +20% | +50% | +50% | -13.8% | +50% | +87.5% | +50% | +13.8% |
| A3 | VR | +13% | +17% | +13% | +17% | +90% | 0% | 0% | 0% | 0% | 0% | 0% | 0% |
|  | OA | +13% | +43% | 0% | +26% | -13.5% | 0% | 0% | 0% | -13.5% | 0% | 0% | +8.3% |
| S1 | VR | +182% | 0% | - - | - - | -100% | 0% | -66.7% | -36% | - - | - - | - - | - - |
|  | OA | -82% | 0% | -82% | 0% | -300% | 0% | +66.7% | 0% | +300% | 0% | +66.7% | +61.9% |
| S2 | VR | -73% | 0% | -33% | +27% | 0% | 0% | 0% | 0% | 0% | 0% | 0% | 0% |
|  | OA | 0% | -27% | -40% | -27% | 0% | 0% | 0% | 0% | 0% | 0% | 0% | 0% |
| S3 | VR | +50% | 0% | -33% | 0% | +50% | 0% | 0% | -10.6% | +50% | 0% | 0% | +14.2% |
|  | OA | +100% | +55% | +133% | +100% | 0% | 0% | 0% | -24.8% | 0% | 0% | 0% | +11.9% |
| S4 | VR | -9% | 0% | -20% | 0% | 0% | 0% | -22% | 0% | -88.7% | 0% | -11% | NM |
|  | OA | 0% | +26% | +43% | +43% | 0% | 0% | +66.7% | 0% | +88.7% | 0% | +88.9% | 0% |
| S5 | VR | +10% | 0% | +8% | 0% | 0% | 0% | 0% | 0% | 0% | 0% | 0% | 0% |
|  | OA | -100% | 0% | 0% | 0% | 0% | 0% | 0% | 0% | 0% | 0% | 0% | 0% |
| M1 | VR | 0% | 0% | 0% | 0% | +50% | +50% | -200% | -204.5% | +60% | +50% | -200% | +41.8% |
|  | OA | -50% | +100% | 0% | +100% | -10% | 0% | -200% | -195.5% | 0% | 0% | -200% | -11.9% |
| M2 | VR | +13% | -30% | - - | - - | +94.4% | 0% | 0% | 0% | - - | - - | - - | - - |
|  | OA | - - | - - | - - | - - | - - | - - | - - | - - | - - | - - | - - | - - |
| M3 | VR | -48% | +20% | -38% | 0% | 0% | 0% | 0% | 0% | 0% | 0% | 0% | 0% |
|  | OA | 0% | 0% | +73% | +13% | 0% | 0% | 0% | 0% | 0% | 0% | 0% | 0% |
| M4 | VR | 0% | +19% | -14% | 0% | +50% | 0% | +88.9% | 0% | 0% | 0% | +88.9% | 0% |
|  | OA | 0% | 0% | 0% | 0% | 0% | 0% | 0% | 0% | 0% | 0% | 0% | 0% |
| M5 | VR | +10% | 0% | 0% | -25% | +77.8% | 0% | 0% | 0% | 0% | 0% | 0% | NM |
|  | OA | 0% | 0% | 0% | 0% | 0% | 0% | 0% | NM | 0% | 0% | 0% | NM |
| M6 | VR | - - | - - | - - | - - | - - | - - | - - | - - | - - | - - | - - | - - |
|  | OA | +27% | 0% | +67% | -10% | -100% | 0% | 0% | +4.2% | 0% | 0% | 0% | +45.8% |

In the OA group, no child improved: 3 (A1, S2, S3) remained stable and 2 (S1, M1) worsened. Among adults, 1 (A2) improved at ST and LT, 5 (S4, S5, M3, M4, M5) remained stable and 2 (A3, M6) deteriorated.

#### 3.2.2. TNO test

Two patients demonstrated measurable stereoacuity at baseline.

In the VR arm, 1 child (M1) improved at ST and LT, while 4 (A1, S1, S2, S3) remained stable. Among adults, no improvement was observed: 1 (A2) deteriorated and 7 remained stable.

In the OA arm, children remained stable. Among adults, 1 (A2) improved at ST and LT, and 7 remained stable.

#### 3.2.3. Lang II test

Four patients showed measurable stereoacuity at baseline.

In the VR arm, no child improved: 3 (A1, S2, S3) remained stable and 2 declined. Among adults, 1 (M4) improved at ST and LT, 5 (A3, S5, M2, M3, M5) remained stable, and 2 (A2, S4) deteriorated.

In the OA arm, 2 children (A1, S1) showed ST improvements, 1 persisting at LT; 2 (S2, S3) remained stable and 1 (M1) worsened. Among adults, 2 (A2, S4) improved at ST and LT, and 6 (A3, S5, M3, M4, M5, M6) remained stable.

#### 3.2.4. Asteroid test

Five patients showed measurable stereoacuity at baseline.

In the VR arm, no child improved: 2 (A1, S2) remained stable and 3 (S1, S3, M1) worsened. Among adults, no improvements were noted: 7 (A3, S4, S5, M2, M3, M4, M5) remained stable and 1 (A2) deteriorated.

In the OA arm, no child improved: 3 (A1, S1, S2) remained stable, 2 (S3, M1) worsened. Among adults, 1 (M6) improved, 5 (A3, S4, S5, M3, M4) remained stable, and 1 (A2) deteriorated at ST, returning to baseline at LT.

### 3.4. Reading speed

One adult (M5) improved in reading speed in the VR arm by 28% in ST.

### 3.5. Compliance

Children had medium compliance (48%), with 1 showing low compliance (18%), 3 medium (34, 53, 64%) and 1 high compliance (73%) (Table 1, two rightwards panels). Adults had high compliance on average (70%), with 1 having low compliance (10%), 2 medium (40 and 47%) and 6 high (70, 78, 89, 97, and 2x100%;).

### 3.6. Adverse events (AE)

Only one AE of transient dizziness while playing was reported by an anisometropic adult (A3).

## 4. Discussion

The AMBER study treated residual amblyopia with binocular, serious, VR-based games. Though designed as an RCT[13], small and heterogeneous cohorts shifted interpretation toward a case-series. Analyzed were BCVA, stereoacuity, reading speed, compliance, and AEs. Data show that VR-based training can improve vision in residual amblyopia patients; however, magnitude and durability of the response varied according to the test, amblyopia subtype, age, and compliance. When comparing patients with each other, VR vs OA, and AmE vs FE, consistency of improvements could not ultimately be confirmed. In detail, BCVA improved in the AmE following VR training in children and adults, at ST and LT. Similar effects have been reported in previous dichoptic or VR-based interventions^9–12,16–18^. Thus, AMBER data support the assumed therapeutic benefit of VR-based video games in treating naïve and residual amblyopia beyond the commonly accepted therapeutic window of 6 years of age. Additionally, AMBER reported data of the OA arm and the FE (supplement), and compared different examination methods, revealing in parts improvements similar to VR-based therapy in both, OA and FE. Nevertheless, there was an unambiguous trend for a higher rate of improvements after VR-based training in AmE (Landolt: 9 VR-based improvement vs. 4x OA-based improvements).

The early emergence of gains in pediatric studies aligns with AMBER, where children improved more despite moderate compliance. Thanks to the study design including children and adults, this age-related difference was detected, highlighting age as a key factor for benefit and emphasizing both the limits of visual plasticity and the importance of early intervention.

When comparing different amblyopia types, BCVA improvements were greatest in anisometropic patients (3/3), followed by strabismic (3/5) and mixed amblyopia patients (2/5), which is consistent with previous studies^11,18^. This result is probably because anisometropic amblyopia primarily affects one eye, which is directly targeted by VR-training, whereas strabismic and mixed types involve more complex binocular deficits that may limit VR training’s efficacy.

Two adults and one child have preserved ST-gained BCVA improvement in LT. This result is consistent with studies from Vedamurthy *et al*. who reported BCVA improvements in adults, with gains at short- and long-term, though long-term gains were weaker and variable^9,10^. Also Gambacorta *et al*. and Manny *et al*. reported well-maintained gains^11,17^. Thus, we should ask why our non-responders had no treatment (long-term) benefits. We assume that treatment has to be customized for different amblyopia types, naïve and residual amblyopia, and age with optimized treatment frequency and duration.

Here, a regimen of 30 min training/day, five days a week for 2 months was applied (20 hours), suggesting immersive VR may allow gains even with low training doses compared to dichoptic action games that may need longer training for positive effects^9,12,17^. Otherwise, Molina-Martin *et al*. reported BCVA improvements in children after only 9 hours of training^12^. Manny *et al*. observed BCVA improvements after 2 and 6 weeks of dichoptic movie viewing in children^17^. Gambacorta *et al*., on the other hand, reported well-maintained gains using a 20-hour protocol comparable to AMBER^11^. Our early emergence of gains in children despite moderate compliance supports the positive association of young age and benefits. Nevertheless, in AMBER (persisting) gain in BCVA was also seen in older highly compliant patients, suggesting that more intense training (longer and/or more frequent) may compensate partially for older age.

Another crucial parameter whose differences make conclusive meta-analyses challenging, are different types of therapy. AMBER tested patients with a mix of dichoptic and binocular VR-based games, as did Meqdad *et al*.^18^. Others used dichoptic action game training^9–11^, 2D home-based iPad games, or watching of dichoptic movies^17^. There seem to be differences in efficiency between different game types, but the role of confounders like age, amblyopia type, and compliance needs to be revealed before the optimal therapy can be defined. Otherwise, it can already be assumed that the common feature, namely, binocular treatment, is generally beneficial.

Different results obtained with the Landolt, and the Sloan chart were notable. Following the Sloan test results, three patients improved, while according to the Landolt test eight patients improved BCVA. The data highlight the importance of selecting carefully tests to be conducted when designing a study and to analyze results of other studies considering the tests performed. Clinically, letter/number optotypes are preferred as they are easier to explain and faster performed ^19,20^. Legally, e.g., in Germany, the Landolt test is used for attestations, assuming a higher sensitivity to small changes when measuring pure-resolution VA, low recognizability, independence of the capability of reading and language, and thus being more reliable and applicable in the whole population^19,20^. Consequently, when compared with each other, BCVA determined with the Landolt test is often lower. However, when analyzing differences in BCVA (like in AMBER), the more challenging test with lower guessing rates, i.e., Landolt, enables the detection of smaller differences, what might explain its more pronounced improvements.

Stereoacuity improved ST in six participants (children and adults). However, only children maintained gains in LT. We interpret this maintenance being related to higher neural plasticity in children. Yet, improvements were only seen in patients with measurable stereovision already at baseline, which is consistent with literature ^11,12,18^. Comparing the different amblyopia types, AMBER patients suffering from mixed amblyopia responded best to VR training (4/6), compared to 1/6 each, anisometropic and strabismic amblyopic patients. This contrasts with two studies that found strong benefits in anisometropic patients, yet the direct comparison with mixed amblyopic patients was missing^9,11^. As suggested by Levi *et al*.^3^, we assume that the immersive 3D VR-training in AMBER, which inherently trains stereopsis, is more effective in improving stereoacuity in mixed amblyopia, compared to dichoptic 2D action games used in previous studies^9,11^.

Most stereovision improvements were detected with tests including monocular cues (Titmus and Lang II), whereas tests minimizing such cues (TNO and Asteroid) classified more participants as stereoblind and detected fewer gains. The former are undoubtedly useful in the clinic and daily life, where compensation strategies play a role, but prove to be unsuitable for quantifying the efficacy of novel therapies.

Reading speed improvements were seen only in one patient. It is assumed that improvements in BCVA were not profound enough to improve this secondary consequence of visual function. More in-depth determination of baseline capabilities and longer follow-up periods could have facilitated detection of such benefits.

VR training was excellently tolerated, with a good mean compliance (62.4%) that is higher than objectively reported patching therapy compliance (40-60%)^21^. It is assumed that binocular video games lead to improved compliance mainly because playing is entertaining, does not suppress using the better eye, and social stigma associated with patching is avoided. Comparably, a review reported a completion of prescribed treatment in >60% of patients in the majority of the 21 analyzed RCTs, yet it is unclear if compliance was subjectively or objectively reported^22^. However, game-based therapy is not inherently successful. Important factors supporting engagement is rendering games challenging but still feasible; this maximizes motivation while minimizing frustration. AMBER used the Vivid Vision games system that adapts games’ difficulty level to visual capacities of the patient in the context of several different short games likely having influenced positively patients’ compliance. Further increases of treatment adherence may be achieved by games that are customized to different age groups. Repka reported poor compliance with a Tetris-like game^23,24^, while a game with animated characters in a gold mine achieved much better compliance (>75% compliance for 68% of patients in 4 weeks)^25^,illustrating how strongly the type of game may influence adherence^23^. Importantly, no AMBER-participant discontinued during the VR intervention. The dropouts occurred either before initiation of treatment or after its completion, suggesting that attrition was not driven from the intervention itself. The home-based design likely facilitated study adherence by reducing practical barriers associated with onsite visits, as seen in other studies with 28% and 38% drop-outs, respectively^9,11^.

Despite thorough study design, AMBER had limitations. The small, heterogeneous sample, due to lower recruitment shortly after the pandemic, prevented statistical analysis of subpopulations and in-depth characterization of different variables (age, amblyopia type). This limited the strength of our conclusions. The two-month-long treatment and follow-up period may have been too short to fully assess responsiveness and durability of improvements.

Summarized, the AMBER study tested home-based, serious, binocular games in immersive VR (20 h play time over 2 months [ST] followed by 2 months of follow-up [LT]) for their efficiency in improving BCVA, stereoacuity, and reading speed in residual amblyopia patients between 9-35 years of age. Improvements were seen in adults and children, at ST and LT, with a trend for stronger and more durable effects with younger age and higher compliance. Yet, a larger RCT should demonstrate results’ consistency. Multi-testing revealed weaknesses of individual tests for use in controlled RCTs, and we recommend the Landolt test to determine BCVA and the Asteroid test to detect stereopsis. Binocular games in VR seem to enable improving vision in all types of residual amblyopia even beyond the traditional therapeutic window. Larger studies with extended treatment regimens are necessary to confirm trends and develop recommendations for efficient, personalized therapy regimens of amblyopia using home-based, serious, binocular games in immersive VR.

## Supporting information

Supplementary Analyses Fellow Eye

STROBE Checklist AMBER study

## Data Availability

Data will be made available on public repositories after finishing and publishing results from all measures.

## Contributions

Conceptualization: PJM, HS, GT, DB, CSM; Methodology: PJM, WB, DB, HS, GT, MK, CSM; Software: BTB, BD, JJB; Validation: PJM, GT, HS, DB, MK, CSM; Formal analysis: AA, NLM, DB, MK, PJM; Investigation: CSM, MPA, WB, HS, PJM; Resources: CSM, HM, GT, HS, PJM; Data curation: AA, NLM, CSM, MPA, DB, MK, PJM; Writing – original draft: AA, NLM, DB, BTB, MK, PJM; Writing – review & editing: AA, NLM, CSM, MPA, WB, DB, BTB, BD, JJB, MK, HM, MM, GT, HS, PJM ; Visualization: AA, NLM, DB, BTB, MK, PJM; Supervision: CSM, WB, MK, GT, HS, PJM; Project administration: CSM, MK, HM, GT, HS, PJM; Funding acquisition: CSM, HM, MK, GT, MM, PJM. All authors approved the final version of the manuscript.

## Institutional Review Board Statement

in text.

## Informed Consent Statement

Informed consent was obtained from all subjects involved in the study.

## Acknowledgments

The authors want to thank Prof. Dennis Levi for his valuable contribution to the conceptualization and analyses of data of the study. We also would like to thank the orthoptists and technicians: Aurore Follonier, Sarah Gisselbaek, and Petra Schampel, for their extraordinary commitment to examining study participants and supporting the result analysis. Thank you also to Lora Fanda for assistance with development of study documents, Alan Purdy and Tuan Tran from Vivid Vision for assistance with the virtual training, and Christian da Fonseca for help with designing the data figures. Preliminary analyses of the study were presented at the Swiss Society of Ophthalmology Annual meeting 2025 and at the Winterthur Ophthalmology Symposium 2026.

## Conflicts of interest

JJB is the CEO and founder of Vivid Vision Inc., and BTB is the Chief Science Officer of Vivid Vision Inc. Vivid Vision Inc. freely provided the licenses to use Vivid Vision Home for the study. Vivid Vision Inc. did not have the ultimate authority on the design, intervention or publication of this study. The other authors declare that they have no competing interests

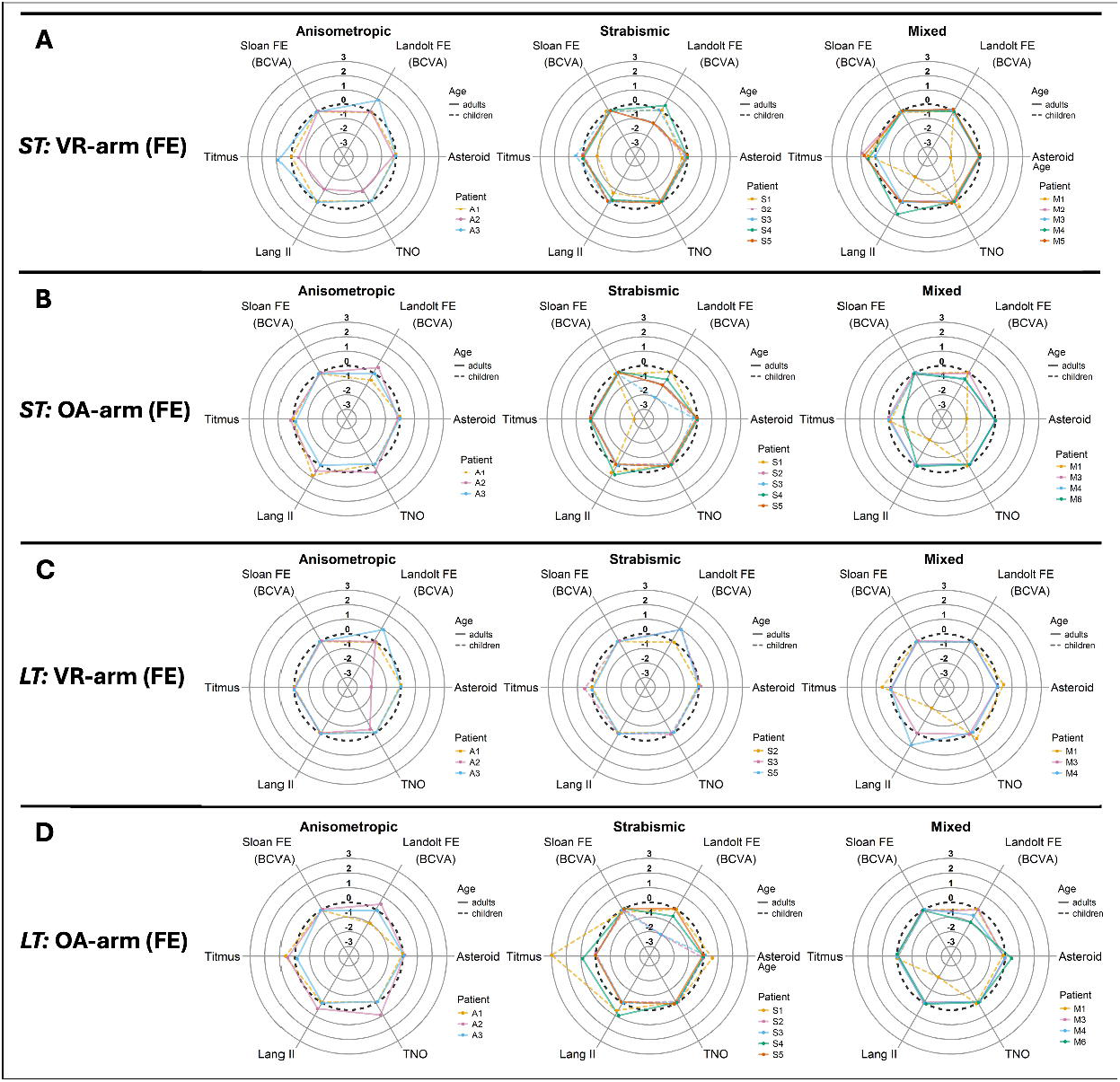

