## Supplementary Analyses Fellow Eye for "Home-based binocular serious games in virtual reality to treat visual acuity and stereovision in residual amblyopia: AMBER study"

*Visual acuity (FE)*

Landolt Chart

In the VR arm, **1 child (S1) improved at ST**; 3 (A1, S3, M1) remained stable at ST, with 1 (S3) improving at LT; and 1 (S2) deteriorated. **Among adults, 3 (A3, S4, M5) improved at ST with 2 (A3, M5) maintaining the gains at LT too**; 4 remained stable (A2, M2, M3, M4); and 1 (S5) deteriorated at ST and LT (Table 1).

In the OA arm, 1 child (S1) improved; 1 (M1) remained stable at ST and LT, and 3 (A1, S2, S3) deteriorated at both ST and LT. Among adults, 2 (A2, M3) improved at both ST and LT; 1 (A3) remained stable; and 5 (S4, S5, M4, M5, M6) deteriorated at both ST and LT, except for 1 (S5) returning to baseline at LT.

Sloan Chart

In the VR arm, **4 children (A1, S2, S3, M1) remained stable, 1 (S1) declined** and all **adults** remained stable (Table 1).

In the OA arm, all children remained stable at ST, with 2 (S1, S2) only improving at LT. All adults remained stable at both ST and LT.

**Table 1. BCVA changes of the fellow eye measured using Landolt and Sloan charts.** Improvements are marked with a “+”, deteriorations with a “-“ and stability with “0%”. The symbol “- - “ indicates that the patient dropped out.

|  |  |  | **ST** | |  |  | **LT** |
| --- | --- | --- | --- | --- | --- | --- | --- |
| **ID** | **Arm** |  | **Landolt** | **Sloan** |  | **Landolt** | **Sloan** |
| A1 | VR |  | 0% | 0% |  | 0% | 0% |
|  | OA |  | -50% | 0% |  | -100% | 0% |
| A2 | VR |  | 0% | 0% |  | 0% | 0% |
|  | OA |  | +50% | 0% |  | +50% | 0% |
| A3 | VR |  | +100% | 0% |  | +100% | 0% |
|  | OA |  | 0% | 0% |  | 0% | 0% |
| S1 | VR |  | +10% | -10% |  | - - | - - |
|  | OA |  | +10% | 0% |  | 0% | +10% |
| S2 | VR |  | -100% | 0% |  | 0% | 0% |
|  | OA |  | -100% | 0% |  | -200% | +10% |
| S3 | VR |  | 0% | 0% |  | +100% | 0% |
|  | OA |  | -200% | 0% |  | -200% | 0% |
| S4 | VR |  | +50% | 0% |  | -50% | 0% |
|  | OA |  | -50% | 0% |  | -50% | 0% |
| S5 | VR |  | -100% | 0% |  | -100% | 0% |
|  | OA |  | -100% | 0% |  | 0% | 0% |
| M1 | VR |  | 0% | 0% |  | 0% | 0% |
|  | OA |  | 0% | 0% |  | 0% | 0% |
| M2 | VR |  | 0% | 0% |  | - - | - - |
|  | OA |  | - - | - - |  | - - | - - |
| M3 | VR |  | 0% | 0% |  | 0% | 0% |
|  | OA |  | +116% | 0% |  | +116% | 0% |
| M4 | VR |  | 0% | 0% |  | 0% | 0% |
|  | OA |  | -50% | 0% |  | -50% | 0% |
| M5 | VR |  | +10% | 0% |  | +10% | 0% |
|  | OA |  | -10% | 0% |  | -10% | 0% |
| M6 | VR |  | - - | - - |  | - - | - - |
|  | OA |  | -50% | 0% |  | -100% | 0% |

**Figure 1. Radar plots showing the effects in BCVA and stereoacuity in the FE at ST and LT, across VR and OA.** Dotted reference line, indicates zero change from baseline; continous line, adults; dashed line, children.

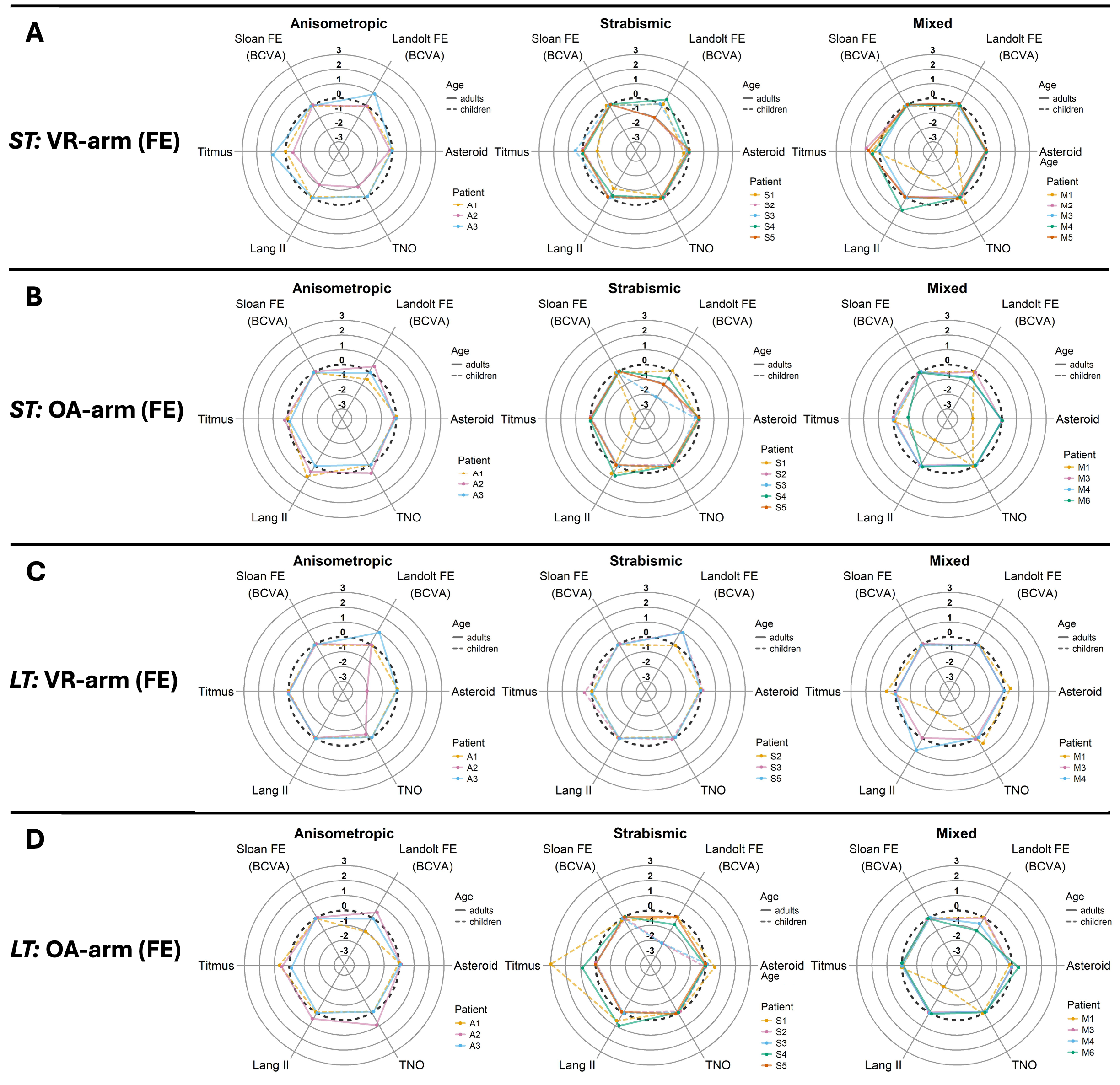
