## Supplementary material for "Home-based binocular serious games in virtual reality to treat visual acuity and stereovision in residual amblyopia: AMBER study": STROBE Checklist AMBER study

STROBE Statement—checklist of items that should be included in reports of observational studies

|  | Item No. | Recommendation | Page  No. | Relevant text from manuscript |
| --- | --- | --- | --- | --- |
| **Title and abstract** | 1 | (*a*) Indicate the study’s design with a commonly used term in the title or the abstract | 1 | “The monocentric, prospective, randomized, crossover trial (reported as case series) included 14 anisometropic, strabismic, or mixed residual amblyopia patients “ |
|  |  | (*b*) Provide in the abstract an informative and balanced summary of what was done and what was found | 1 | “Methods and Analysis” and “Results” sections of Abstract. |
| Introduction | | | |  |
| Background/rationale | 2 | Explain the scientific background and rationale for the investigation being reported | 2 | “Introduction” section, 1^st^ paragraph |
| Objectives | 3 | State specific objectives, including any prespecified hypotheses | 3 | “Present prospective, single-center, randomized crossover clinical trial (RCT) evaluated the effectiveness of binocular stimulation using eight complementary serious VR-based games as an adjunct to optical correction (OA) in patients with residual amblyopia of age beyond the therapeutic window (protocol paper published^13^). The comparison of multiple BCVA and stereopsis tests aimed at identifying differences and finding the most appropriate (combination of) methods to be used. We hypothesized that VR-based games would improve BCVA, stereovision, reading, attentional and motor skills in patients with residual amblyopia, regardless of age.” |
| Methods | | | |  |
| Study design | 4 | Present key elements of study design early in the paper | 3 | “AMBER was a prospective, blinded, monocentric, cross-over RCT, conducted in accordance with Declaration of Helsinki & approved by the Ethical Cantonal commission of Geneva (N° 2021-D0090, approved 8.11.22); the limited number and heterogeneity of enrolled patients led us report the results as case series.” |
| Setting | 5 | Describe the setting, locations, and relevant dates, including periods of recruitment, exposure, follow-up, and data collection | 3 | “Nineteen patients were recruited between 05/2022 and 12/2024. “; “…and examinations were performed at the Department of Ophthalmology, University Hospitals of Geneva. “; “The VR arm involved eight binocular serious games embedded in a VR headset, allowing home-based training after an introduction at the clinic (Vivid Vision, San Francisco, USA). Games were played, while wearing prescribed glasses, for 30 min/day limited to this maximum duration by automatic locking, 5 days/week, over 2 months.” |
| Participants | 6 | (*a*) *Cohort study*—Give the eligibility criteria, and the sources and methods of selection of participants. Describe methods of follow-up  *Case-control study*—Give the eligibility criteria, and the sources and methods of case ascertainment and control selection. Give the rationale for the choice of cases and controls  *Cross-sectional study*—Give the eligibility criteria, and the sources and methods of selection of participants | 3 | “Eligible participants were aged 6-35 years with residual amblyopia (anisometropic, strabismic or mixed), defined as BCVA < 20/20 in the amblyopic eye (AmE) and an interocular difference of ≥2 lines with refractive correction. BCVA had to be stable across at least 2 consecutive assessments obtained ≥6 months after the last treatment.”; “Nineteen patients were recruited between 05/2022 and 12/2024 from among patients at HUG Opthalmology.”; “Two certified, orthoptists performed ocular examinations during 5 visits scheduled per patient; another team member assessed non-ocular outcomes. All (and other two members, analyzing the data) were blinded. Visits took place at baseline (T1, screening), after the first intervention (T2, short-term [ST]), 2 months after the end of the first intervention (T3, follow-up 1, long-term [LT] and screening for the second intervention), after the second intervention (T4, [ST]), and after the second follow-up (T5, [LT])” |
|  |  | (*b*) *Cohort study*—For matched studies, give matching criteria and number of exposed and unexposed  *Case-control study*—For matched studies, give matching criteria and the number of controls per case | 3 | Study designed as a cross-over RCT thus each patient were their own control : “Patients (nonblinded) completed two 2-month-long interventions in a randomized order: OA (optical correction alone) (patients wore glasses with prescribed correction), and VR (patients played binocular serious video games in addition to OA).” |
| Variables | 7 | Clearly define all outcomes, exposures, predictors, potential confounders, and effect modifiers. Give diagnostic criteria, if applicable | 3 | Outcomes: Sections “2.4. Primary outcome” and “2.5. Secondary outcome”  Residual amblyopia : “with residual amblyopia (anisometropic, strabismic or mixed), defined as BCVA < 20/20 in the amblyopic eye (AmE) and an interocular difference of ≥2 lines with refractive correction. BCVA had to be stable across at least 2 consecutive assessments obtained ≥6 months after the last treatment.” |
| Data sources/ measurement | 8* | For each variable of interest, give sources of data and details of methods of assessment (measurement). Describe comparability of assessment methods if there is more than one group | 4 | Sections “2.4. Primary outcome” and “2.5. Secondary outcome” |
| Bias | 9 | Describe any efforts to address potential sources of bias | 3 | “All (and other two, analyzing the data) were blinded.” |
| Study size | 10 | Explain how the study size was arrived at | 5 | “Target sample of 30 participants, powered for primary outcome^13^...” |

Continued on next page

| Quantitative variables | 11 | Explain how quantitative variables were handled in the analyses. If applicable, describe which groupings were chosen and why | 5 | “The small, heterogeneous sample led us analyze data descriptively (min, max, SD, SEM, range), reporting individual patient outcomes as detailed case studies. If not otherwise mentioned, data are reported as mean±SD, separately for adults and children, and 3 amblyopia subtypes.” |
| --- | --- | --- | --- | --- |
| Statistical methods | 12 | (*a*) Describe all statistical methods, including those used to control for confounding | 5 | As just above |
|  |  | (*b*) Describe any methods used to examine subgroups and interactions | 5 | As just above (“If not otherwise mentioned, data are reported as mean±SD, separately for adults and children, and 3 amblyopia subtypes.”) |
|  |  | (*c*) Explain how missing data were addressed | 3 | “Ultimately, 14 patients were evaluated (data of all are reported here),” |
|  |  | (*d*) *Cohort study*—If applicable, explain how loss to follow-up was addressed  *Case-control study*—If applicable, explain how matching of cases and controls was addressed  *Cross-sectional study*—If applicable, describe analytical methods taking account of sampling strategy |  | Study designed as a cross-over RCT thus each patient were their own control. Follow-up data loss reported in paper. |
|  |  | (*e*) Describe any sensitivity analyses |  | N/A |
| Results | | | | |
| Participants | 13* | (a) Report numbers of individuals at each stage of study—eg numbers potentially eligible, examined for eligibility, confirmed eligible, included in the study, completing follow-up, and analysed | 3 | “Nineteen patients were recruited between 05/2022 and 12/2024 at HUG Opthalmology. Two patients did not meet inclusion criteria, three dropped out after T1. Ultimately, 14 patients were evaluated (data of all are reported here), with 13 completing both interventions.” |
|  |  | (b) Give reasons for non-participation at each stage | 3 | As just above |
|  |  | (c) Consider use of a flow diagram | 14 | Figure 1 |
| Descriptive data | 14* | (a) Give characteristics of study participants (eg demographic, clinical, social) and information on exposures and potential confounders | 5 | Section 3.1 Sample characteristics |
|  |  | (b) Indicate number of participants with missing data for each variable of interest | 5 | Section 3.1 Sample characteristics ; “All BCVA and stereoacuity measures had 4 and 2 missing data, for VR and OA, respectively (12 of these ST, 24 LT).” |
|  |  | (c) *Cohort study*—Summarise follow-up time (eg, average and total amount) |  | 2 months post each treatment arm, for all participants |
| Outcome data | 15* | *Cohort study*—Report numbers of outcome events or summary measures over time |  |  |
|  |  | *Case-control study—*Report numbers in each exposure category, or summary measures of exposure | 5 | “All 14 patients attended T1 and T2. At T3, 1 patient dropped out (n = 13), another before T4 (n = 12), and a third one by T5 (n = 11). (…) All BCVA and stereoacuity measures had 4 and 2 missing data, for VR and OA, respectively (12 of these ST, 24 LT).” |
|  |  | *Cross-sectional study—*Report numbers of outcome events or summary measures |  |  |
| Main results | 16 | (*a*) Give unadjusted estimates and, if applicable, confounder-adjusted estimates and their precision (eg, 95% confidence interval). Make clear which confounders were adjusted for and why they were included | 5 | N/A “The small, heterogeneous sample led us analyze data descriptively (min, max, SD, SEM, range), reporting individual patient outcomes as detailed case studies.” |
|  |  | (*b*) Report category boundaries when continuous variables were categorized | 4,5 | Only Asteroid test and MNRead are continuous scales , respectively “Every possible level within the measurable range (13-1,200 arcsec) is assessed.” and “Recorded metrics were the smallest print size read without errors, the smallest print size read at maximum speed, the maximum reading speed measured in words per minute (wpm). “ |
|  |  | (*c*) If relevant, consider translating estimates of relative risk into absolute risk for a meaningful time period |  | N/A |

Continued on next page

| Other analyses | 17 | Report other analyses done—eg analyses of subgroups and interactions, and sensitivity analyses | 5 | A study designed as an RCT but reported as a series of detailed cases; “The small, heterogeneous sample led us analyze data descriptively (min, max, SD, SEM, range), reporting individual patient outcomes as detailed case studies. If not otherwise mentioned, data are reported as mean±SD, separately for adults and children, and 3 amblyopia subtypes.” |
| --- | --- | --- | --- | --- |
| Discussion | | | | |
| Key results | 18 | Summarise key results with reference to study objectives | 8 | “Data show that VR-based training can improve vision in residual amblyopia patients; however, magnitude and durability of the response varied according to the test, amblyopia subtype, age, and compliance. When comparing patients with each other, VR vs OA, and AmE vs FE, consistency of improvements could not ultimately be confirmed. In detail, BCVA improved in the AmE following VR training in children and adults, at ST and LT. Similar effects have been reported in previous dichoptic or VR-based interventions^9–12,16–18^. Thus, AMBER data support the assumed therapeutic benefit of VR-based video games in treating naïve and residual amblyopia beyond the commonly accepted therapeutic window of 6 years of age. Additionally, AMBER reported data of the OA arm and the FE (supplement), and compared different examination methods, revealing in parts improvements similar to VR-based therapy in both, OA and FE. Nevertheless, there was an unambiguous trend for a higher rate of improvements after VR-based training in AmE (Landolt: 9 VR-based improvement vs. 4x OA-based improvements).” |
| Limitations | 19 | Discuss limitations of the study, taking into account sources of potential bias or imprecision. Discuss both direction and magnitude of any potential bias | 10 | “Despite thorough study design, AMBER had limitations. The small, heterogeneous sample, due to lower recruitment shortly after the pandemic, prevented statistical analysis of subpopulations and in-depth characterization of different variables (age, amblyopia type). This limited the strength of our conclusions. The two-month-long treatment and follow-up period may have been too short to fully assess responsiveness and durability of improvements. “ |
| Interpretation | 20 | Give a cautious overall interpretation of results considering objectives, limitations, multiplicity of analyses, results from similar studies, and other relevant evidence | 9 | “Thus, AMBER data support the assumed therapeutic benefit of VR-based video games in treating naïve and residual amblyopia beyond the commonly accepted therapeutic window of 6 years of age.” (…)When comparing different amblyopia types, BCVA improvements were greatest in anisometropic patients (3/3), followed by strabismic (3/5) and mixed amblyopia patients (2/5), which is consistent with previous studies^11,18^. This result is probably because anisometropic amblyopia primarily affects one eye, which is directly targeted by VR-training, whereas strabismic and mixed types involve more complex binocular deficits that may limit VR training’s efficacy.  Two adults and one child have preserved ST-gained BCVA improvement in LT. This result is consistent with studies from Vedamurthy *et al.* who reported BCVA improvements in adults, with gains at short- and long-term, though long-term gains were weaker and variable^9,10^. Also Gambacorta *et al.* and Manny *et al*. reported well-maintained gains^11,17^. Thus, we should ask why our non-responders had no treatment (long-term) benefits. We assume that treatment has to be customized for different amblyopia types, naïve and residual amblyopia, and age with optimized treatment frequency and duration.  Here, a regimen of 30 min training/day, five days a week for 2 months was applied (20 hours), suggesting immersive VR may allow gains even with low training doses compared to dichoptic action games that may need longer training for positive effects^9,12,17^. Otherwise, Molina-Martin *et al.* reported BCVA improvements in children after only 9 hours of training^12^. Manny *et al.* observed BCVA improvements after 2 and 6 weeks of dichoptic movie viewing in children^17^. Gambacorta *et al.*, on the other hand, reported well-maintained gains using a 20-hour protocol comparable to AMBER^11^. Our early emergence of gains in children despite moderate compliance supports the positive association of young age and benefits. Nevertheless, in AMBER (persisting) gain in BCVA was also seen in older highly compliant patients, suggesting that more intense training (longer and/or more frequent) may compensate partially for older age.  “ |
| Generalisability | 21 | Discuss the generalisability (external validity) of the study results | 10 | “Larger studies with extended treatment regimens are necessary to confirm trends and develop recommendations for efficient, personalized therapy regimens of amblyopia using home-based, serious, binocular games in immersive VR.” |
| Other information | |  | | |
| Funding | 22 | Give the source of funding and the role of the funders for the present study and, if applicable, for the original study on which the present article is based | 10 | Section “Funding” |

*Give information separately for cases and controls in case-control studies and, if applicable, for exposed and unexposed groups in cohort and cross-sectional studies.

**Note:** An Explanation and Elaboration article discusses each checklist item and gives methodological background and published examples of transparent reporting. The STROBE checklist is best used in conjunction with this article (freely available on the Web sites of PLoS Medicine at http://www.plosmedicine.org/, Annals of Internal Medicine at http://www.annals.org/, and Epidemiology at http://www.epidem.com/). Information on the STROBE Initiative is available at www.strobe-statement.org.
